# Uganda Genome Resource: A rich research database for genomic studies of communicable and non-communicable diseases in Africa

**DOI:** 10.1101/2022.05.05.22274740

**Authors:** Segun Fatumo, Joseph Mugisha, Opeyemi S Soremekun, Allan Kalungi, Richard Mayanja, Christopher Kintu, Ronald Makanga, Ayoub Kakande, Andrew Abaasa, Gershim Asiki, Robert Kalyesubula, Robert Newton, Moffat Nyirenda, Manj S. Sandhu, Pontiano Kaleebu

**Affiliations:** The African Computational Genomics (TACG) Research Group, MRC/UVRI and LSHTM, Entebbe, Uganda; London School of Hygiene and Tropical Medicine London, United Kingdom; Medical Research Council/ Uganda Virus Research Institute/London School of Hygiene and Tropical Medicine (MRC/UVRI/LSHTM) Uganda research unit, Entebbe, Uganda; College of Health Sciences, Makerere University, Kampala Uganda; Health and Systems for Health Research Unit, African Population and Health Research Center, Nairobi, Kenya; Department of Epidemiology and Biostatistics, School of Public Health, Imperial College London, United Kingdom

**Keywords:** Genomics, Uganda, NCDs, GPC

## Abstract

The Uganda Genome Resource (UGR) is a well characterised genomic database, with a range of phenotypic communicable and non-communicable diseases and risk factors generated from the Uganda General Population Cohort (GPC) - a population-based open cohort study established in 1989 by the Medical Research Council (MRC) UK in collaboration with the Uganda Virus Research Institute (UVRI).

In 2011, UGR was launched with genotype data on ∼5000 and whole genome sequence data on ∼2000 Ugandan individuals from 9 ethno-linguistic groups. Leveraging other available platforms at the MRC Uganda such as Biorepository centre for sample storage, Clinical Diagnostic Laboratory Service (CDLS) for sample diagnostic testing, sequencing platform for DNA extraction, Uganda Medical informatics Unit (UMIC) for large-scale data analysis, GPC for additional sample collection, UGR is strategically poised to expand and generate scientific discoveries.

Here, we describe UGR and highlight the important genetic findings thus far including how UGR is providing opportunities to: (1) discover novel disease susceptibility genetic loci; (2) refine association signals at new and existing loci; (3) develop and test Polygenic Risk Score (PRS) to determine individual’s disease risk; 4) assess how some risk factors including infectious diseases are causally related to non-communicable diseases (NCDs) in Africa; (5) develop research capacity for genomics in Africa; and (6) enhance African participation in the global genomics research arena. Leveraging established research infrastructure, expertise, local genomic leadership, global collaboration and strategic funding, we anticipate that UGR can develop further to a comparable level of European and Asian large-scale genomic initiatives.

## Introduction

The genetic diversity in Africa is far greater than in any other region across the globe but unfortunately, the vast majority of genomic studies have been performed in European ancestry populations (PMID: 35145307). Uganda is located in East Africa with four major ethic groups and over 40 languages. The rich linguistic, ethnic, and cultural diversity of Uganda provides an unprecedented opportunity to understand the level of the genetic structure in Uganda populations. To advance genetic epidemiology of communicable and non-communicable diseases (NCDs) in Uganda, the Ugandan Genome Resource (UGR) was launched in 2011 to prospectively collect a wide range of NCDs, infectious disease risk factors including information on lifestyle, family history social determinant, demographics, sexual health & reproductive behaviour, past illness, mental health, treatment and immunisation and environmental risk factors (Asiki *et al*., 2013).

Here, we provide a detailed description of UGR which is different from previous publications on GPC that focused on specific aspects (Asiki *et al*., 2013) or population genetics and genome-wide association analyses of cardiometabolic traits in UGR data (Gurdasani *et al*., 2019), we aim to give an overview of UGR as a resource including detailed phenotype availability, genomic data generation, sample characteristics, genetic discoveries to date, and finally to its data access and sharing policy.

**Figure 1:**
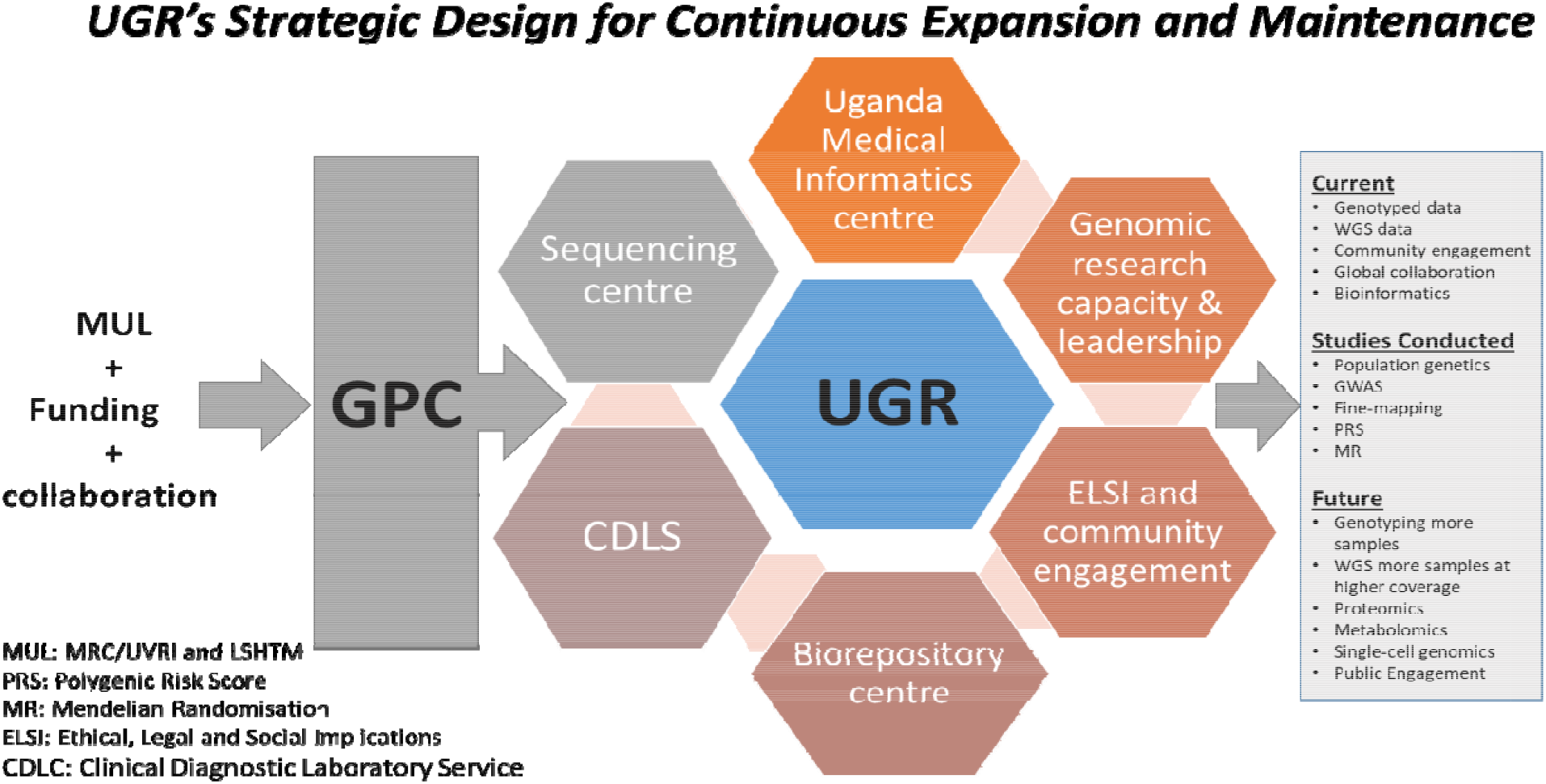
UGR strategic vision for continuous expansion and maintenance. UGR continues to offer as a valuable platform for investigating the genetics of NCDs and relationship with infectious diseases in Africa.

### Study Population – The General Population Cohort (GPC)

The GPC is a population-based study of approximately 22,000 individuals residing in 25 neighbouring villages in the Kyamulibwa sub-county, Kalungu district in rural south-western Uganda. The study was founded in 1989 by the Medical Research Council UK (MRC UK) in collaboration with the Uganda Virus Research Institute (UVRI) to study the epidemiology of HIV in a general population. The GPC population was initially recruited and assessed through annual house-to-house census and survey rounds until 2012, when biannual surveys commenced. Since its establishment, 26 rounds of survey and 29 rounds of census have been undertaken. Before any survey procedures are carried out, written informed consent is obtained from participants on the use of their clinical records for research purposes and sample storage for future use (Asiki et al., 2013). Data collected includes serological, demographic, and medical information from participants. Information regarding mortality, fertility, sexual behaviour migration, HIV infection perception are routinely collated.

The GPC Round 22 study of 2011 focused on the genetics and epidemiology of communicable and non-communicable disease. The survey round which was used to establish the Ugandan Genome Resource (UGR) consisted of five main stages, including mobilization (recruitment and consenting), mapping, census, survey, and results feedback and clinical follow-up. The specific objectives of this survey then were:

- To create a one-of-a-kind study for expanding on a large-scale prospective cohort research in an African population to evaluate a wide range of health indices—and to lay the platform for longer-term investigations.
- Using population, genetic and epidemiological techniques to provide aetiological insights into variance in cardio-metabolic and infectious risk factors.
- To help develop public health policies in other African countries by informing health policy and public health programs aimed at addressing the rise in NCDs in Uganda.

The cohort continues to offer as a valuable platform for investigating the relationship between communicable illnesses and NCDs in a regular annual survey of GPC.

### Genotype generation, quality control and Imputation

The 2.5M Illumina chip array was used to genotype nearly 5,000 Ugandans at the Wellcome Trust Sanger Institute. Gurdasani and Fatumo et al 2019 have presented the quality control steps (Gurdasani *et al*., 2019). In summary, we used a strict quality control process to perform a series of steps in a logical order to eliminate a total of 39,368 autosomal markers that failed to meet the quality metrics for SNP call rate (>97 percent, 25,037 SNPs) and HWE (p<1×10^−8^, 14,331 SNPs). During sample QC, a total of 91 samples were eliminated because they failed the quality standards for sample call rate (>97%) or heterozygosity (HO=0.209333±0.007416 matching to the mean±3SD), or the sex extrapolated from the X-chromosome did not correspond to the reported sex. Three further samples were eliminated due to high relatedness (IBD>0.90). There were no samples that were classified as outliers in terms of population or ancestry. 2,230,258 autosomal markers and 4778 samples that met the stated threshold were subjected to further analysis. We carried out SNP Phasing with the aid of SHAPEIT2 (Delaneau, Coulonges and Zagury, 2008) using default settings, then imputation was done with IMPUTE2 (Howie, Donnelly and Marchini, 2009). All samples were imputed with a combined reference that was created by combining the UG2G sequence resource (n = 2,000, whole genome sequence data from the African Genome Variation Project (n = 320),), and the 1000 Genomes phase 3 project (n = 2,504). The principal components analysis plot for the GPC participants (n=4,778) was published (Gurdasani et al., 2019) and is shown here in supplementary material figure S1.

### Uganda 2000 Genomes (UG2G)

The entire genomes of over 2,000 Ugandans from nine ethnolinguistic groups were sequenced using the Illumina HiSeq 2000 with 75bp paired end reads at low coverage, with an average coverage of 4x for each sample. 343 of these samples overlapped with people who had already been genotyped. An automated quality control process was used to bring down the data files that needs manual processing to ascertain the quality of BAM files produced. This method was based on the one developed for the UK10K project (Walter *et al*., 2015) which used a set of algorithmically derived standards to determine summary data computed from the input BAMs. Any line that fell below the “fail” standard for any of the metrics was deleted; while lines falling below the warn standard for any of the scores were manually investigated; and any line that passes any of these scores was given a status of “pass”. Overall, we deleted fourteen samples from the study. Full detailed on the quality control and how we computed the summary data has be described in Gurdasani *et al*., 2019.

### Merging of Sequenced and Genotyped Data

We integrated sequenced and imputed genotyped data to produce an aggregated dataset to boost power for discovery in a genome-wide association studies. Because cryptic and family relatedness persisted across sequenced and genotyped data, we produced an aggregated dataset for analysis instead of separately meta-analysing the data, because data would be correlated rather than independent. As a result, conclusions from mixed model analysis that explicitly model this relationship are more likely to be true. We examined and deleted any consistent discrepancies between sequences and imputed genotype data after merging the two datasets. This was done by performing principal component analysis on the dataset to see if there was any distinction by data modality (imputed genotype data vs. sequenced data) among the 343 people who had their genotypes and sequences done in duplicate. On PCA, we noticed a strong separation of genotype imputed and sequence data points. For these 343 samples, we tested alternative concordance criteria between sequencing and imputed genotype data, screening out SNPs with a concordance of 0.80 and 0.90 the dataset. We discovered that to eliminate systematic effects detected between genotyping array and sequence data on PCA, a minimum concordance criterion of 0.90 was necessary.

There were no systematic changes between sequenced and genotyped data in PCAs after excluding 904,283 SNPs that exhibited 90 percent consonance in genotypes between the sequence and imputed genotype data. We examined the top ten PCs to confirm that systematic variations in the genomic data did not constitute an important axis of variation.

### Phenotype and laboratory measurement

During survey round 22 which was conducted in 2011, several phenotypes based on clinical and physical examinations, laboratory tests, and self-reported questionnaires were collected from the respondents (Table 1) and these respondents who are still known to be alive and have not moved out of the GPC have been followed every year since then. A blood specimen was analysed for non-fasting blood lipids, blood cell traits (mean cell haemoglobin, red cell count, white cell count, mean cell haemoglobin concentration, haemoglobin, packed cell volume, mean cell volume and platelet), glycaemic characteristics, renal function, infectious biomarkers (HIV, hepatitis B and C). Basic demographics such as age, sex, marital status, and education level; anthropometrics such as BMI, weight, waist-to-hip ratio, height; blood pressure measurements; and lifestyle information such as smoking status, physical activities, and diet; sexual health & reproductive behaviour; sex education, condom use, pregnancy & outcome, and number of offspring were also collected (Table 1).

**Table 1:** Sample characteristics of UGR participants at baseline

| **Characteristic** | **All**<br>**(n=7833)** | **Males**<br>**(n=3425)** | **Female**<br>**(n=4404)** |
| --- | --- | --- | --- |
| Number of individuals interviewed, N | 7833 | 3425 | 4404 |
| Age (years), median (IQR) | 30 (17-46) | 27 (17-44) | 31 (19-47) |
| **Age group (years), N(%)** |  |  |  |
| 13-19 | 2398 (30.6) | 1218 (35.6) | 1180 (26.8) |
| 20-29 | 1475 (18.8) | 612 (17.9) | 863 (19.6) |
| 30-39 | 1311 (16.8) | 495 (14.5) | 816 (18.5) |
| 40-49 | 1047 (13.4) | 439 (12.8) | 608 (13.8) |
| 50-59 | 711 (9.1) | 297 (8.7) | 414 (9.4) |
| 60+ | 887 (11.3) | 364 (10.6) | 523 (11.9) |
| BMI (kg/m<sup>2</sup>), mean ± SD | 21.2 ± 3.83 | 20.1 ± 3.11 | 22.0 ± 4.13 |
| **BMI Classification, N(%)** |  |  |  |
| Underweight | 1712 (22.6) | 1031 (30.4) | 681 (16.3) |
| Normal | 4919 (65.0) | 2188 (64.4) | 2731 (65.4) |
| Overweight | 739 (9.8) | 156 (4.6) | 583 (14.0) |
| Obese | 201 (2.6) | 21 (0.6) | 180 (4.3) |
| **Smoking Status, N(%)** |  |  |  |
| Current Smoker | 641 (8.2) | 553 (16.2) | 88 (2.0) |
| Ex-Smoker | 194 (2.5) | 169 (4.9) | 25 (0.6) |
| Never smoked | 6990 (89.3) | 2700 (78.9) | 4290 (97.4) |
| **Alcohol Consumption, N(%)** |  |  |  |
| Never or no alcohol use | 5040 (70.1) | 2052 (64.6) | 2988 (74.4) |
| Infrequent current drinker | 537 (7.5) | 165 (5.2) | 372 (9.3) |
| Frequent current drinker | 1618 (22.5) | 961 (30.2) | 657 (16.4) |
| **Cardio metabolic Quantitative measurements (mean ± SD)** |  |  |  |
| TC (mmol/L), mean ± SD | 3.5 ± 0.98 | 3.3 ± 0.91 | 3.7 ± 1.00 |
| HDL (mmol/L), mean ± SD | 1.0 ± 0.41 | 0.9 ± 0.42 | 1.0 ± 0.40 |
| LDL (mmol/L), mean ± SD | 2.0 ± 0.77 | 1.8 ± 0.63 | 2.1 ± 0.80 |
| Albumin, mean ± SD | 41.3 ± 4.10 | 41.8 ± 4.15 | 40.9 ± 4.02 |
| HbA1c, mean ± SD | 3.3 ± 0.68 | 3.3 ± 0.69 | 3.3 ± 0.73 |
| TG (mmol/L), mean ± SD | 1.2 ± 0.61 | 1.1 ± 0.61 | 1.2 ± 0.62 |
| ALT, median (IQR) | 14.0 (17.8-22.9) | 19.4 (15.6-25.1) | 13.0 (16.4-21.3) |
| ALK, median (IQR) | 71.3 (92.5-144.1) | 74.3 (96.8-208.0) | 68.5 (89.5-123.1) |
| AST, median (IQR) | 21.2 (25.1-30.4) | 23.8 (28.0-33.0) | 19.8 (23.1-27.4) |
| GGT, median (IQR) | 13.5 (18.7-28.0) | 15.6 (21.6-33.0) | 12.2 (17.0-24.2) |
| Bilirubin, median (IQR) | 5.2 (7.7-12.0) | 5.92 (8.9-14.2) | 4.8 (6.9-10.5) |
| **Anthropometric Measurements** |  |  |  |
| Weight (kg), mean ± SD | 52.6 ± 11.35 | 52.4 ± 11.37 | 52.7 ± 11.33 |
| Height (cm) , mean ± SD | 157.2 ± 9.19 | 160.7 ± 10.54 | 154.5 ± 6.83 |
| SBP (mmHg) , mean ± SD | 122.4 ± 17.0 | 123.5 ± 16.2 | 121.6 ± 17.51 |
| DBP (mmHg) , mean ± SD | 74.2 ± 10.26 | 73.5 ± 10.39 | 74.7 ± 10.12 |
| **Anaemia** |  |  |  |
| WBC | 5.1 ± 1.51 | 5.1 ± 1.58 | 5.1 ± 1.58 |
| RBC | 4.7 ± 0.62 | 4.9 ± 0.65 | 4.6 ± 0.56 |
| HGB | 13.6 ± 1.62 | 14.2 ± 1.74 | 13.1 ± 1.33 |
| WHR | 0.85 ± 0.16 | 0.86 ± 0.17 | 0.8 ± 0.16 |
| MCH | 28.9 ± 2.91 | 29.1 ± 2.92 | 28.8 ± 2.90 |
| MCHC | 33.7 ± 1.19 | 33.7 ± 1.24 | 33.7 ± 1.15 |
| RDW | 13.1 ± 1.34 | 13.1 ± 1.40 | 13.1 ± 1.29 |
| MPV | 8.7 ± 0.83 | 8.7 ± 0.83 | 8.7 ± 0.82 |
| Platelet count (PLT) | 216.9 ± 77.7 | 207.9 ± 77.3 | 223.9 ± 77.30 |
| Lymphocytes | 2.4 ± 0.83 | 2.5 ± 0.92 | 2.4 ± 0.76 |
| Monocytes | 0.3 ± 0.12 | 0.29 ± 0.14 | 0.3 ± 0.11 |
| Basophils | 0.05 ± 0.04 | 0.05 ± 0.42 | 0.05 ± 1.58 |
| Neutrophils | 1.9 ± 0.86 | 1.9 ± 0.84 | 2.0 ± 0.88 |
| Eosinophils | 0.35 ± 0.39 | 0.4 ± 0.40 | 0.3 ± 0.39 |
| Vaccination |  |  |  |
| **Received BCG vaccine, N(%)** |  |  |  |
| Yes | 1421 (18.2) | 631 (18.4) | 790 (18.0) |
| No | 553 (7.1) | 221 (6.5) | 332 (7.5) |
| Don't know | 5850 (74.8) | 2570 (75.1) | 3280 (74.5) |
| **Received Oral Polio vaccine, N(%)** |  |  |  |
| Yes | 1398 (17.9) | 612 (17.9) | 786 (17.9) |
| No | 578 (7.4) | 237 (6.9) | 341 (7.8) |
| Don't know | 5848 (74.7) | 2573 (75.2) | 3275 (74.7) |
| **Received DPT vaccine, N(%)** |  |  |  |
| Yes | 1415 (18.1) | 626 (18.3) | 789 (17.9) |
| No | 541 (6.9) | 212 (6.2) | 329 (7.5) |
| Don't know | 5868 (75.0) | 2584 (75.5) | 3284 (74.6) |
| **Received Measles vaccine, N(%)** |  |  |  |
| Yes | 1561 (20.0) | 685 (20.0) | 876 (19.9) |
| No | 561 (7.2) | 226 (6.6) | 335 (7.6) |
| Don't know | 5702 (72.9) | 2511 (73.4) | 3191 (72.5) |
| **Received TB vaccine, N(%)** |  |  |  |
| Yes | 54 (0.7) | 22 (0.6) | 32 (0.7) |
| No | 7326 (92.5) | 3131 (91.5) | 4105 (93.2) |
| Don't know | 535 (6.8) | 269 (7.9) | 266 (6.1) |
| **Received Hepatitis B vaccine, N(%)** |  |  |  |
| Yes | 54 (0.69) | 22 (0.6) | 32 (0.7) |
| No | 7258 (92.8) | 3141 (91.8) | 4117 (93.5) |
| Don't know | 513 (6.56) | 259 (7.6) | 254 (5.8) |
| **Received Tetanus vaccine, N(%)** |  |  |  |
| Yes | 1881 (24.0) | 9 (0.3) | 1872 (42.5) |
| No | 5462 (69.8) | 3154 (92.2) | 2308 (52.4) |
| Don't know | 482 (6.2) | 259 (7.6) | 223 (5.1) |
| **Received Tetanus booster vaccine, N(%)** |  |  |  |
| Yes | 1285 (16.4) | 298 (8.7) | 987 (22.4) |
| No | 6044 (77.3) | 2863 (83.7) | 3181 (72.3) |
| Don't know | 495 (6.3) | 260 (7.6) | 235 (5.3) |
| **Received Rabies vaccine, N(%)** |  |  |  |
| Yes | 46 (0.6) | 17 (0.5) | 29 (0.7) |
| No | 7268 (92.9) | 3138 (91.7) | 4130 (93.8) |
| Don't know | 511 (6.5) | 257 (7.8) | 244 (5.5) |
| **Self-report of diseases** |  |  |  |
| **Hypertension, N(%)** |  |  |  |
| Hypertensive | 487 (6.2) | 130 (3.8) | 357 (8.1) |
| Normal | 7338 (93.8) | 3292 (96.2) | 4046 (91.9) |
| **Diabetes, N (%)** |  |  |  |
| Diabetic | 102 (1.3) | 44 (1.3) | 58 (1.3) |
| Normal | 7723 (98.7) | 3378 (98.7) | 4345 (98.7) |
| **Blood test results** |  |  |  |
| **HIV status, N (%)** |  |  |  |
| Negative | 7185 (92.4) | 3197 (94.0) | 3988 (91.1) |
| Positive | 593 (7.6) | 204 (6.0) | 389 (8.9) |
| **Hepatitis B, N (%)** |  |  |  |
| Negative | 7536 (97.3) | 3268 (96.5) | 4268 (97.9) |
| Positive | 210 (2.7) | 117 (3.5) | 93 (2.1) |
| **Hepatitis C, N (%)** |  |  |  |
| Negative | 7536 (97.3) | 3268 (96.5) | 4268 (97.9) |
| Positive | 210 (2.7) | 117 (3.5) | 93 (2.1) |

### Genetic discoveries and polygenic prediction in UGR

A case in point in the use of our rich African genomics and phenotypic data, we undertook GWAS in 34 cardiometabolic traits including lipid, anthropometry traits, blood cell indices, HbA1c and reported novel loci associated with anthropometric, haematological, lipid, and glycaemic traits (Gurdasani *et al*., 2019). In another study (Fatumo et al 2020 - eGFR), we reported the first ever GWAS of kidney function in continental Africa. Leveraging clinical relatedness and correlations among phenotypes, we explored the power of multivariate GWAS to identify genetic risk factors implicating pleiotropic effects in blood cell traits (Fatumo *et al*., 2019; Soremekun *et al*., 2021), body shape (Nakabuye *et al*., 2022) and liver function. Recently, we showed that genetic risk score derived from data of African American individuals enhance polygenic prediction of lipid traits and T2DM in Sub-Sahara African, but prediction varied greatly between another dataset from South Africa and our East African genomic data (Chikowore *et al*., 2022, Kamiza *et al*., 2022). We have also demonstrated the Mendelian randomisation evidence of relation between lipid trait and T2DM (Soremekun *et al*., 2022), metabolic traits and stroke (Fatumo *et al*., 2021). Collectively, our studies show a need for improved representation of Africans in genomic studies and ensuring the generalisation of findings for genomic medicine. This is further supported by findings from another study as well (Martin et al., 2017). The UGR data has also been used to create a genotype imputation reference panel using UG2G available from the Sanger Imputation Service (imputation.sanger.ac.uk).

### Contribution to collaborative studies

We contribute to global genetic studies through partnerships and consortia, such as the African Partnership for Chronic Disease Research (APCDR), an international network of research groups that collaborate to support and promote collaborative chronic disease research across Africa. An initiative created in response to the changing distribution of communicable diseases and the rising burden of noncommunicable diseases, as well as the recognition that low- and middle-income countries (LMICs), including those in Sub-Saharan Africa, will need to expand their health-care capacities to effectively respond to these epidemiological transitions.

We combine research expertise with three other MRC Units (MRC Integrative Epidemiology Unit, MRC Population Health Research Unit, and MRC Unit for Lifelong Health and Ageing) to investigate the potential to use Mendelian randomization (MR) to assess the generalisability of existing drugs (e.g., statins, anti-diabetics, and anti-hypertensives) and identify the potential to tailor drugs with pilot studies focusing on established pharmacological targets to specific subpopulations (e.g. *CETP, HMGCR*) and to see how changes in genetic architecture affect efficacy estimates in different groups.

We are part of the CARDINAL (CARDiometabolic Disorders IN African-ancestry PopuLations) which is study site of an NIH-funded Polygenic Risk Methods in Diverse Populations (PRIMED) Consortium https://primedconsortium.org/. CARDINAL (Adebamowo, C.A. *et al*., 2022) aims to integrate phenotype and genomic datasets from 50,000 African individuals from seven cohort studies and evaluate PRSs to develop a novel method that considers ancestry-specific genomic regions to improve PRS prediction in populations with genetic substructure.

Furthermore, we recently provided GWAS data to the Meta-Analyses of Glucose and Insulin-related Variables Consortium (MAGIC) in order to find additional loci that influence glycaemic and metabolic traits (Chen, J. et al 2021). We are aiming for opportunities to contribute key phenotypes such as lipids, blood cell traits, kidney, etc to other consortia. For GBMI we will contribute all phenotype in Table 1 when require, including opportunity to measure not previously collected phenotype using resources in our organisation. We believe that team science allows scientists to make the most progress toward breakthrough discoveries that benefit human health.

### Future directions

The GPC is an active cohort of more than 22,000 participants. Genotype and sequence data is available for 6,657 respondents (N=5,000, 2,000 and 343 for genotype, sequence and overlapping samples respectively). We hope to genotype more samples to add on this resource. We also hope to sequence more samples at higher coverage in order to provide a reference panel with increased genome coverage. We also hope to extend our research into proteomics, metabolomics and single cell genomics in order to gain insights into the different mechanisms and pathways that could be implicated in different disease processes.

### Data access and sharing of the UGR data

Request for resources and information should be directed to UGR’s Data Access Committee (DAC) via the Lead Contact, Dr. Segun Fatumo. UGR’s individual level data, genotype and sequence data are available under managed access to researchers. Requests for access will be granted for all research consistent with the consent provided by participants. This would include any research in the context of health and disease, that does not involve identifying the participants in any way.

The array and low and high depth sequence data have been deposited at the European Genome-phenome Archive (EGA, https://www.ebi.ac.uk/ega/, accession numbers EGAS00001001558/EGAD00010000965, EGAS00001000545/EGAD00001001639 and EGAS00001000545/EGAD00001005346 respectively. Requests for access to data may be directed to. Applications are reviewed by data access committee (DAC) and access is granted if the request is consistent with the consent provided by participants. The data producers may be consulted by the DAC to evaluate potential ethical conflicts. Requestors also sign an agreement which governs the terms on which access to data is granted.

However, full GWAS summary statistics of UGR is freely available on GWAS catalog https://www.ebi.ac.uk/gwas/ with study accssion numbers: GCST009041 (Eosinophil counts), GCST009042 (Total cholesterol levels), GCST009043 (LDL cholesterol levels), GCST0090414 (HDL cholesterol levels), GCST009045 (Triglyceride levels), GCST009046(Aspartate aminotransferase levels), GCST009047 (Alanine aminotransferase levels), GCST009048 (Serum albumin levels), GCST009049 (Serum alkaline phosphatase levels), GCST009050 (Gamma glutamyl transferase levels), GCST009051 (Bilirubin levels), GCST009052 (Diastolic blood pressure) GCST009053 (Systolic blood pressure), GCST009054 (Hemoglobin A1c levels), GCST009055 (Height), GCST009056 (Weight), GCST009057 (Body mass index), GCST009058 (Waist circumference), GCST009059 (Hip circumference), GCST009060 (Waist-hip ratio), GCST009061 (White blood cell count), GCST009062 (Red blood cell count), GCST009063 (mean corpuscular hemoglobin), GCST009064 (mean corpuscular hemoglobin concentration), GCST009065 (mean corpuscular volume), GCST009032 (red blood cell distribution width), GCST009033 (hematocrit), GCST009034 (hemoglobin measurement), GCST009035 (mean platelet volume), GCST009036 (platelet count), GCST009037 (lymphocyte count), GCST009038 (monocyte count), GCST009039 (basophil count), GCST009040 (neutrophil count)

## Conclusions

The Uganda Genome Resource is designed to make direct impact in biomedical and genetic research of health and disease in Uganda, Africa and globally. UGR has become one of the model genomic resources in Africa and offers training opportunities to researchers from Uganda and the world at large. Here we present an overview of the UGR, showcase its broad range of phenotypic data, and highlights the genetic discoveries from UGR till date. In the next few years, UGR will continue to grow in sample size, and include proteomics, metabolomics, and single-cell genomic studies.

## Data Availability

Request for resources and information should be directed to UGR Data Access Committee (DAC) via the Lead Contact Dr. Segun Fatumo. UGR individual level data, genotype and sequence data are available under managed access to researchers. Requests for access will be granted for all research consistent with the consent provided by participants. This would include any research in the context of health and disease, that does not involve identifying the participants in any way.

## Ethics

The study was approved by the Science and Ethics Committee of the Uganda Virus Research Institute Research (UVRI) and Ethics Committee (UVRI-REC #HS 1978) and the Uganda National Council for Science and Technology (UNCST #SS 4283) and the East of England-Cambridge South (formerly Cambridgeshire 4) NHS Research Ethics Committee UK SD, standard deviation; TC, Total Cholesterol; LDL, Low-density lipoprotein; TG, Triglycerides; HDL, High-density lipoprotein; ALT, Alanine aminotransferase; AST, Aspartate aminotransferase; ALP, Alkaline phosphatase; GGT, Gamma glutamyltransferase; DBP, diastolic blood pressure; SBP, systolic blood pressure; BMI, Body mass index; WHR, Waist-Hip Ratio; WBC, White blood cell, RBC, Red blood cell, MCV, mean corpuscular volume; MCH, mean corpuscular haemoglobin; MCHC, mean corpuscular hemoglobin concentration; RDW, red blood cell distribution width; DPT, Diphtheria, pertussis and tetanus vaccine

## Supplementary material

**Figure S1:**
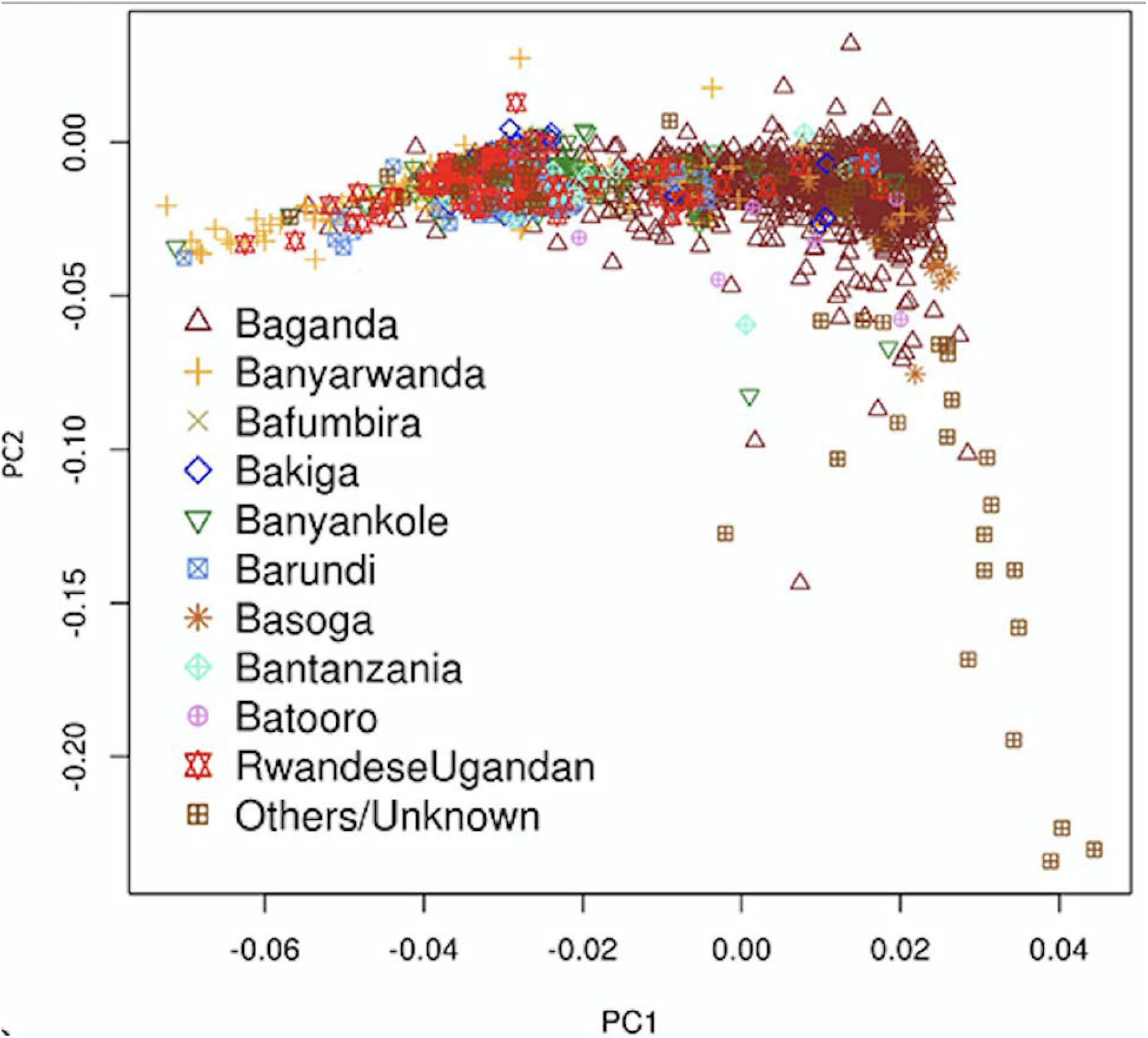
The principal components analysis plot for the general population cohort participants (Source: Gurdasani D *et al*., 2019)

